# The influence of genetic predisposition and physical activity on risk of Gestational Diabetes Mellitus in the nuMoM2b cohort

**DOI:** 10.1101/2022.03.08.22271868

**Authors:** Kymberleigh A. Pagel, Hoyin Chu, Rashika Ramola, Rafael F. Guerrero, Judith H. Chung, Samuel Parry, Uma M. Reddy, Robert M. Silver, Jonathan G. Steller, Lynn M. Yee, Ronald J. Wapner, Matthew W. Hahn, Sriraam Natarajan, David M. Haas, Predrag Radivojac

## Abstract

**Importance:** Polygenic risk scores (PRS) for Type II Diabetes Mellitus (T2DM) can improve risk prediction for Gestational Diabetes Mellitus (GDM), yet the strength of the relationship between genetic and lifestyle risk factors has not been quantified.

**Objective:** To assess the effects of PRS and physical activity on existing GDM risk models and identify patient subgroups who may receive the most benefits from receiving a PRS or activity intervention.

**Design, Settings, and Participants:** The Nulliparous Pregnancy Outcomes Study: Monitoring Mothers-to-Be (nuMoM2b) study was established to study individuals without previous pregnancy lasting 20 weeks or more (nulliparous) and to elucidate factors associated with adverse pregnancy outcomes. A sub-cohort of 3,533 participants with European ancestry were used for risk assessment and performance evaluation.

**Exposures:** Self-reported total physical activity in early pregnancy was quantified as metabolic equivalent of tasks (METs) in hours/week. Polygenic risk scores were calculated for T2DM using contributions of 85 single nucleotide variants, weighted by their association in the DIAbetes Genetics Replication And Meta-analysis (DIAGRAM) Consortium data.

**Main Outcomes and Measures:** Prediction of the development of GDM from clinical, genetic, and environmental variables collected in early pregnancy. The risk model is assessed using measures of model discrimination and calibration. Odds ratio and positive likelihood ratio were used for evaluating the effect of PRS and physical activity on GDM risk.

**Results:** In high-risk population subgroups (body mass index ≥ 25 or age ≥ 35), individuals with PRS in the top 25^th^ percentile or METs below 450 have significantly increased odds of GDM diagnosis. Participants with both high PRS and low METs have three times higher odds of GDM diagnosis than the population. Conversely, participants with high PRS and METs ≥ 450 do not exhibit increased odds of GDM diagnosis, and those with low METs and low PRS have reduced odds of GDM. The relationship between PRS and METs was found to be nonadditive.

**Conclusions and Relevance:** In high-risk patient subgroups the addition of PRS resulted in increased risk of GDM diagnosis, suggesting the benefits of targeted PRS ascertainment to encourage early intervention. Increased physical activity is associated with decreased risk of GDM, particularly among individuals genetically predisposed to T2DM.

**Key Points:** *Question:* Do genetic predisposition to diabetes and physical activity in early pregnancy cooperatively impact risk of Gestational Diabetes Mellitus (GDM) among nulliparas?

*Findings:* Risk of GDM diagnosis increases significantly for nulliparas with high polygenic risk score (PRS) and with low physical activity. The odds ratio of developing GDM with high PRS was estimated to be 2.2, 1.6 with low physical activity, and 3.5 in combination.

*Meaning:* Physical activity in early pregnancy is associated with reduced risk of GDM and reversal of excess risk in genetically predisposed individuals. The interaction between PRS and physical activity may identify subjects for targeted interventions.

## Introduction

Every year approximately 7% of pregnancies in the United States are affected by Gestational Diabetes Mellitus (GDM),^1^ and the risk for developing Type II Diabetes Mellitus (T2DM) has doubled in the past decade among patients with GDM.^2^ GDM has been shown to increase the longer-term risk of maternal T2DM, cardiovascular morbidity, and renal disease, as well as to increase the risk for T2DM, obesity, and neuropsychiatric morbidity in offspring.^3^ To reduce the incidence of GDM and accompanying adverse perinatal outcomes, there remains a need to implement effective and targeted risk reduction strategies among high-risk individuals.

The strongest predictors for the development of GDM are adverse maternal outcomes in previous pregnancies.^4,5^ However, this information is unavailable for 40% of all pregnancies in the U.S. that occur in nulliparous women (nulliparas).^6^ Additional characteristics associated with elevated risk of GDM include obesity, low physical activity, age over 35 years, and family history of diabetes.^7-11^ Despite these insights, the prediction of adverse perinatal outcomes in first pregnancies has remained challenging, and the search for additional GDM risk factors applicable to nulliparas remains ongoing.^12^

Significant progress has been made in identifying genetic variants associated with T2DM and GDM.^13^ Studies incorporating polygenic risk score (PRS) generated from using loci associated with T2DM showed modest improvement in performance for GDM prediction when compared to a baseline clinical model.^14^ However, the added value of physical activity to these models has not been assessed. Moreover, the relationship between PRS and physical activity on the incidence of GDM also remains understudied. Here, using the Nulliparous Pregnancy Outcomes Study: Monitoring Mothers-to-Be (nuMoM2b) observational cohort study of nulliparas,^15^ we provide evidence of the impact of exercise and PRS—both individually and jointly—on the risk of GDM.

## Methods

### Study Population

The study population was composed of individuals who were enrolled in an observational nuMoM2b cohort study where nulliparous women were recruited from hospitals affiliated with eight clinical sites in the United States.^15^ Participants were enrolled from 2010-2013 and four study visits were scheduled at 6-13, 16-21, 22-29 weeks’ gestation, and at the time of the delivery.

There were 10,038 participants in the nuMoM2b cohort. Samples were selected for this work based upon clinical covariates and incidence of GDM (Figure 1). Individuals missing all covariates (*n* = 10), those with pre-diabetes (*n* = 151), those with diagnosis or treatment for diabetes before pregnancy (*n* = 42), and those who were not tested for GDM (*n* = 467) were excluded from the analysis. Of the remaining 9,368 participants, those who were not self-reported White (*n* = 3,702), whose ancestry was not inferred to be European using SNPweights^16^ (*n* = 632), and those with incomplete or erroneous physical exercise data (*n* = 1,214) were excluded. The remaining 3,533 participants (132 GDM cases; 3.7%) were included for further analysis.

**Figure 1.**
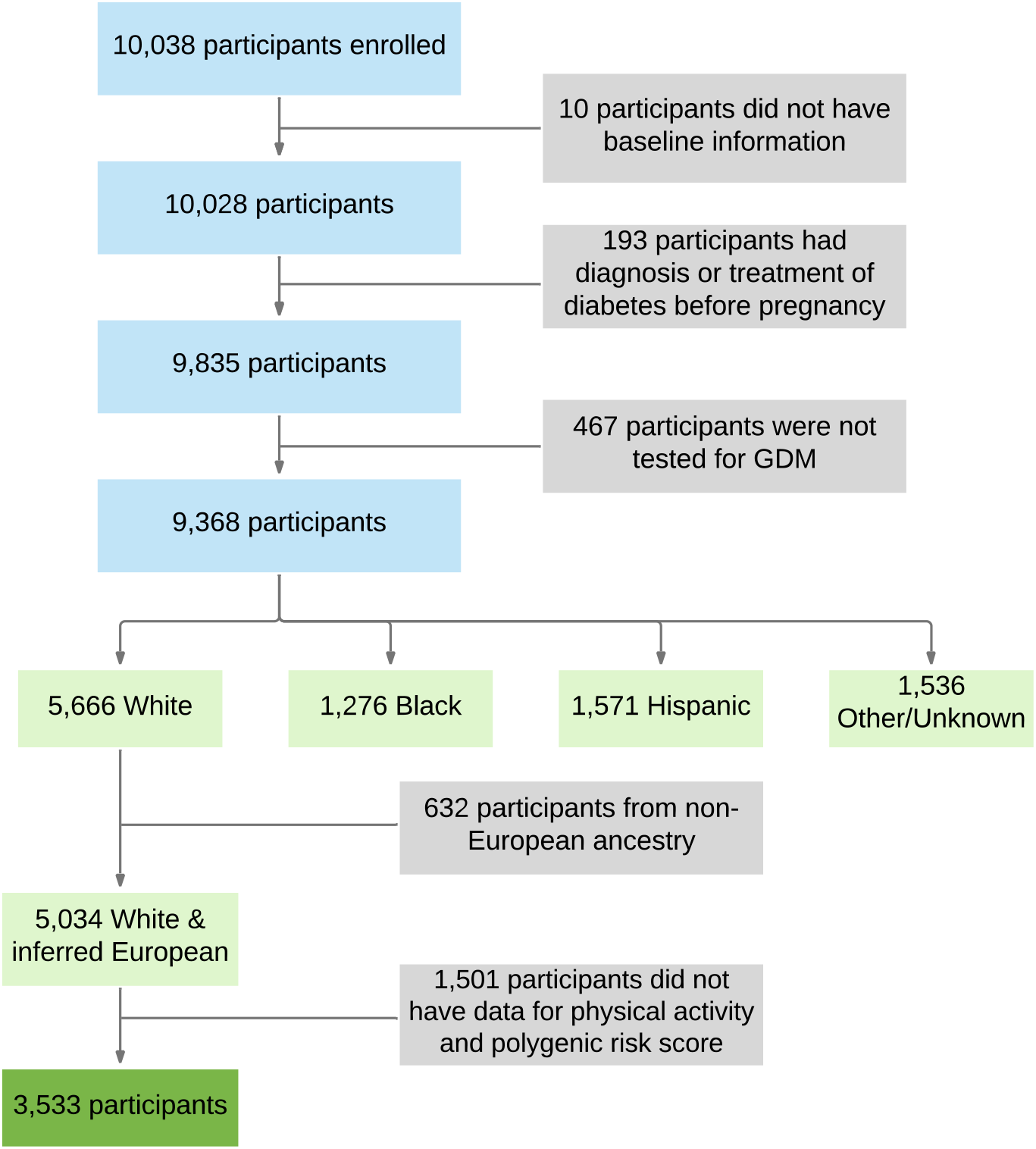
Data preprocessing flowchart. The filtering criteria used to generate the final cohort for analysis.

### Covariates

Data were collected for the nuMoM2b cohort through interviews, self-administered questionnaires, clinical measurements, and medical records. Pregnancy outcome information was collected from medical records, and maternal blood was collected for DNA.^17^ Information on leisure physical activities during pregnancy was reported at study visits 1-3 using questions adapted from the Behavior Risk Factor Surveillance System (https://www.cdc.gov/brfss). Participants’ physical activity during the previous four weeks was reported as the number of times per week and duration in minutes for their most frequently performed activities. The activities were translated to total metabolic equivalent of tasks (METs), calculated as the weighted sum of the minutes spent in each physical activity.^18^ For simplicity, we refer to METs-minutes as METs here onwards. Consistent with the Nurses’ Health Study II (NHS II),^19^ the participants were categorized into less active (*n* = 1,168) and more active (*n* = 2,365) groups based on a METs threshold of 450, which is equivalent to 150 minutes of moderate-intensity or 75 minutes of vigorous-intensity physical activity per week. All time-varying covariates used in this work, including body mass index (BMI), waist circumference, and physical activity data, pertain to the first study visit.

### Processing of Biological Samples and Genotyping Platform

The genotyped cohort comprised participants who had adequate samples and agreed to be genotyped. DNA extractions from whole blood were carried out on a QIAsymphony instrument at the Center for Genomics and Bioinformatics, Indiana University, Bloomington. Genotyping was completed at the Van Andel Institute (Grand Rapids, MI) using the Infinium Multi-Ethnic Global D2 BeadChip (Illumina, USA). Genotype calls for the 1,748,280 loci that passed initial quality control were made with Beeline AutoConvert (Illumina, USA). To estimate genotyping error rates and to increase quality control, we genotyped and replicated a subset of 48 of the individuals for whom we had duplicate blood samples. Using this pilot batch, we verified the quality of our genotyping pipeline (yielding >99.6% call rate in all samples) and estimated a baseline error rate of 6.6×10^−5^ per marker, based on unfiltered single nucleotide variants (SNVs) and using the standard intensity clusters provided by Illumina.^16^

### Calculation of Genetic Risk

Calculation of polygenic risk was performed using summary statistics identified in a genome-wide association study (GWAS) from European populations, as studies based upon non-European populations were not available at the time of analysis. Genotype imputation was done using the pre-phasing and imputation stepwise approach implemented in Eagle and Minimac3, with variable chunk size of 132 genomic chunks and default parameters, against the 1000 Genomes Phase-3 reference.^20^ PRS for T2DM were calculated as in Powe et al.^21^ The method included contributions of 85 SNVs^22^ weighted by the beta coefficients from the DIAGRAM Consortium data.^23^

### Primary Outcome

Gestational diabetes was determined based on one of the following glucose tolerance testing (GTT) criteria: (i) fasting 3-hour 100g GTT with two abnormal values: fasting ≥95 mg/dL, 1-hour ≥180 mg/dL, 2-hour ≥155 mg/dL, 3-hour ≥140 mg/dL; (ii) fasting 2-hour 75g GTT with one abnormal value: fasting ≥92 mg/dL, 1-hour ≥180 mg/dL, 2-hour ≥153 mg/dL; (iii) non-fasting 50g GTT ≥200 mg/dL if no fasting 3-hour or 2-hour GTT was performed. In addition to the GTT data, it was noted if a diagnosis of GDM was made during clinical care. In the absence of GTT data, the clinical diagnosis was used to determine GDM. All outcomes were collected by certified medical record abstractors at each clinical site to make a final determination of GDM diagnosis.^15^

### Models and Evaluation

We first developed a baseline model by using features taken from the list of screening questions obtained from Artzi et al.^4^ The features include age, BMI, race, diabetes history in the family, polycystic ovary syndrome diagnosis, high blood pressure diagnosis, diabetes diagnosis, previous history of GDM, and previous HbA1c% test. Adapting the baseline model to features available to nulliparas in nuMoMs2b, we omitted prior pregnancy history and HbA1c% test from the feature list and incorporated waist size as an additional feature. The performance levels of classification models were measured using areas under the Receiving Operating Characteristic curve (AUCs),^24^ estimated via 10-fold cross-validation. The 95% confidence intervals were determined using 100 bootstrapping iterations.

The test for interaction between covariates was carried out using the following model

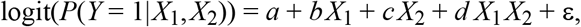

where *Y* is a binary response representing GDM diagnosis (*Y* = 1 indicates presence of GDM), *X*_1_ and *X*_2_ are binary random variables representing high PRS and low METs, respectively, ε is a zero-mean Gaussian random variable with unknown variance σ^2^ and (*a, b, c, d*) is a set of real-valued parameters that, along with σ^2^, were estimated from data. The p-value for coefficient *d* (null hypothesis: *d* = 0) was used to demonstrate nonadditive relationship between *X*_1_ and *X*_2_.

### Calculation of Odds Ratios, Likelihood Ratios and Statistical Significance

Unless otherwise stated, the odds ratio (OR)^25^ of each subgroup was calculated using the rest of the cohort who were not in the subgroup as the reference group, and p-values were obtained using Fisher’s exact test. Positive likelihood ratio (LR^+^)^25^ values were calculated as the ratio of posterior to prior odds of GDM diagnosis, where the prior odds refer to the odds of GDM diagnosis in a parent group and the posterior odds refer to the odds of GDM diagnosis in a subgroup of the parent group; that is, the parent group with additional criteria imposed. LR^+^ p-values were obtained by calculating the proportion of samples in which the child subgroup had a lower (or higher, as appropriate) likelihood ratio than the parent subgroup using 10,000 bootstrapped samples of the analysis cohort.^26^

## Results

Participants diagnosed with GDM exhibited higher T2DM PRS in comparison to controls (Figure 2A; *p*<0.001, t-test), with a prominent excess of cases in the highest score quartile. Participants with high PRS were associated with an increased risk of GDM, particularly when the odds ratio (OR) exceeded 2.5 (Figure 2B). Higher physical activity was associated with a reduced risk of GDM (Figure 2C; *p* = 0.003, t-test). Participants diagnosed with GDM exhibited significantly lower physical activity in early pregnancy compared to participants who were not diagnosed with GDM, with those in the 4^th^ quartile having OR below 0.5 (Figure 2D).

**Figure 2.**
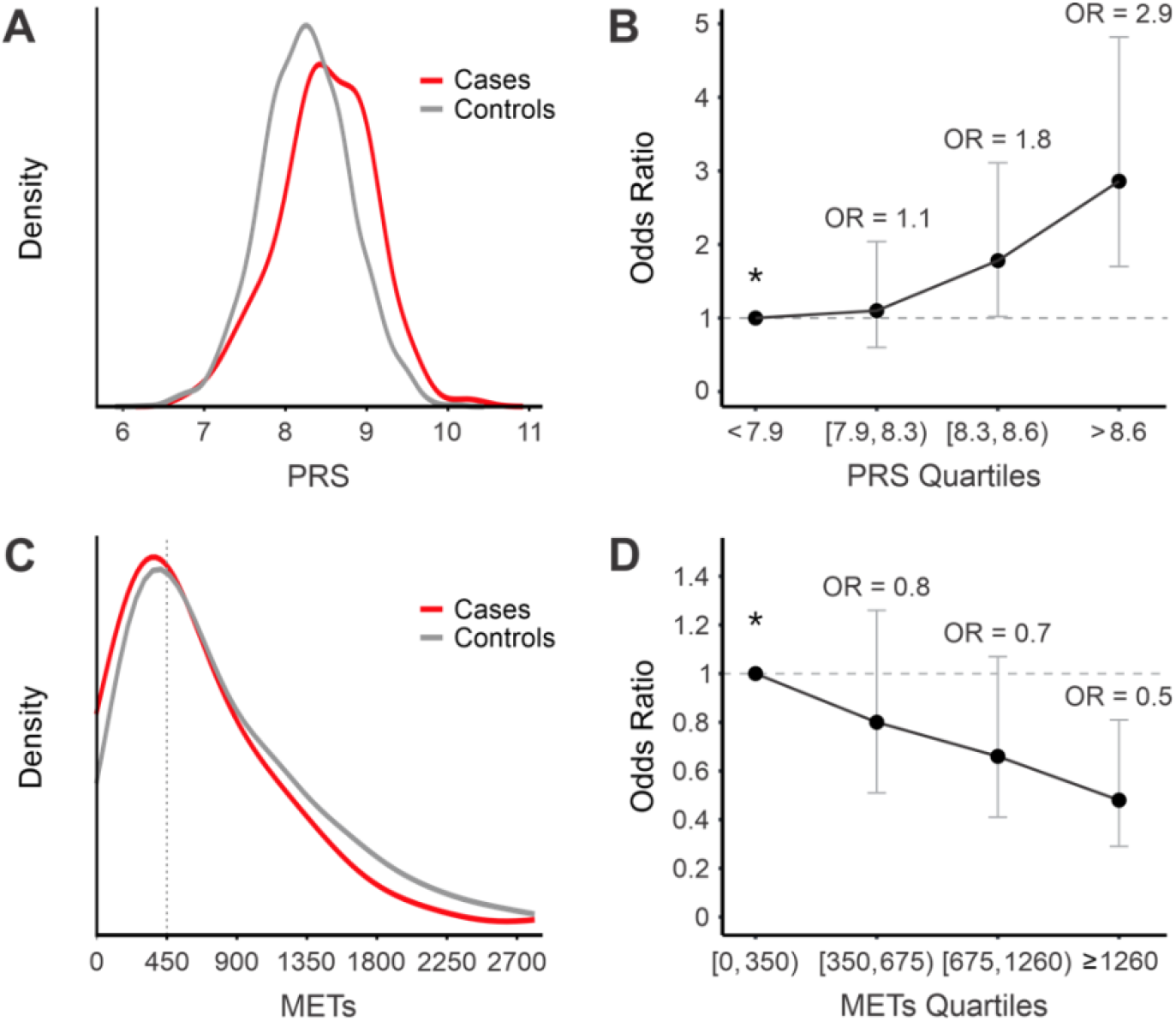
Influence of PRS and METs on GDM risk. (A) PRS Distribution. The distributions of PRS between participants who developed GDM (Cases, red lines) and those who did not (Controls, gray lines). (B) PRS OR Quartiles with 95% confidence intervals. Participants were divided into quartiles based on their PRS, with OR calculated against the reference group Q_1_, denoted by an asterisk. The quartile ORs are OR = 1.1 (0.6, 2.0) for Q_2_, OR = 1.8 (1.0, 3.1) for Q_3_, and OR = 2.9 (1.7, 4.8) for Q_4_. (C) METs Distribution. The distributions of METs between GDM cases and controls. (D) METs OR Quartiles with 95% confidence intervals. Participants were divided into four separate groups based on their METs, with OR calculated against the reference group Q_1_, denoted by an asterisk. The quartile ORs are OR = 0.8 (0.5, 1.3) for Q_2_, OR = 0.7 (0.4, 1.1) for Q_3_, and OR = 0.5 (0.3, 0.8) for Q_4_. Kernel density plots in panels A and C were generated using the kdeplot function (Python Seaborn library v.0.11.2) with default Gaussian kernel. Each density was normalized independently with the argument common_norm set to False.

We next established the influence of PRS and METs on the risk of GDM when participants were stratified based on three high-risk covariates: family history of diabetes, age ≥ 35 years, and BMI ≥ 25 (Figure 3). Participants with family history of diabetes and high PRS, defined here as those with scores above the top 25^th^ percentile, exhibited increased odds of GDM diagnosis compared to the remainder of participants (OR = 2.6). Similarly, participants with family history of diabetes that reported lower levels of physical activity (METs<450) reflected increased odds for GDM compared to the remainder of the cohort (OR = 3.3). Among the participants with family history of diabetes (LR^+^ = 2.0), those with high PRS, had slightly increased odds of GDM diagnosis but the result was not statistically significant (LR^+^ = 2.4, *p*\* = 0.369). On the other hand, those who also reported low levels of METs showed a larger and statistically significant increase in odds for GDM diagnosis than the parent group (LR^+^ = 2.9; *p*\* = 0.015). A similar outcome was observed for the subgroups with low PRS, defined as those with scores below the 25^th^ percentile, and those with higher levels of physical activity (METs≥450). That is, among participants with family history of diabetes, low PRS was not associated with significantly lower odds for GDM (LR^+^ = 1.8; *p*\* = 0.369), whereas high physical activity provided significant reduction in odds for GDM among these participants (LR^+^ = 1.4; *p*\* = 0.016). This suggests that, for the patients with family history of diabetes, information about physical activity may be more informative than PRS. The METs threshold of 450 from the NHS II^19^ was determined to be a well-selected value based upon our exploratory analysis (Supplementary Figure S1).

**Figure 3.**
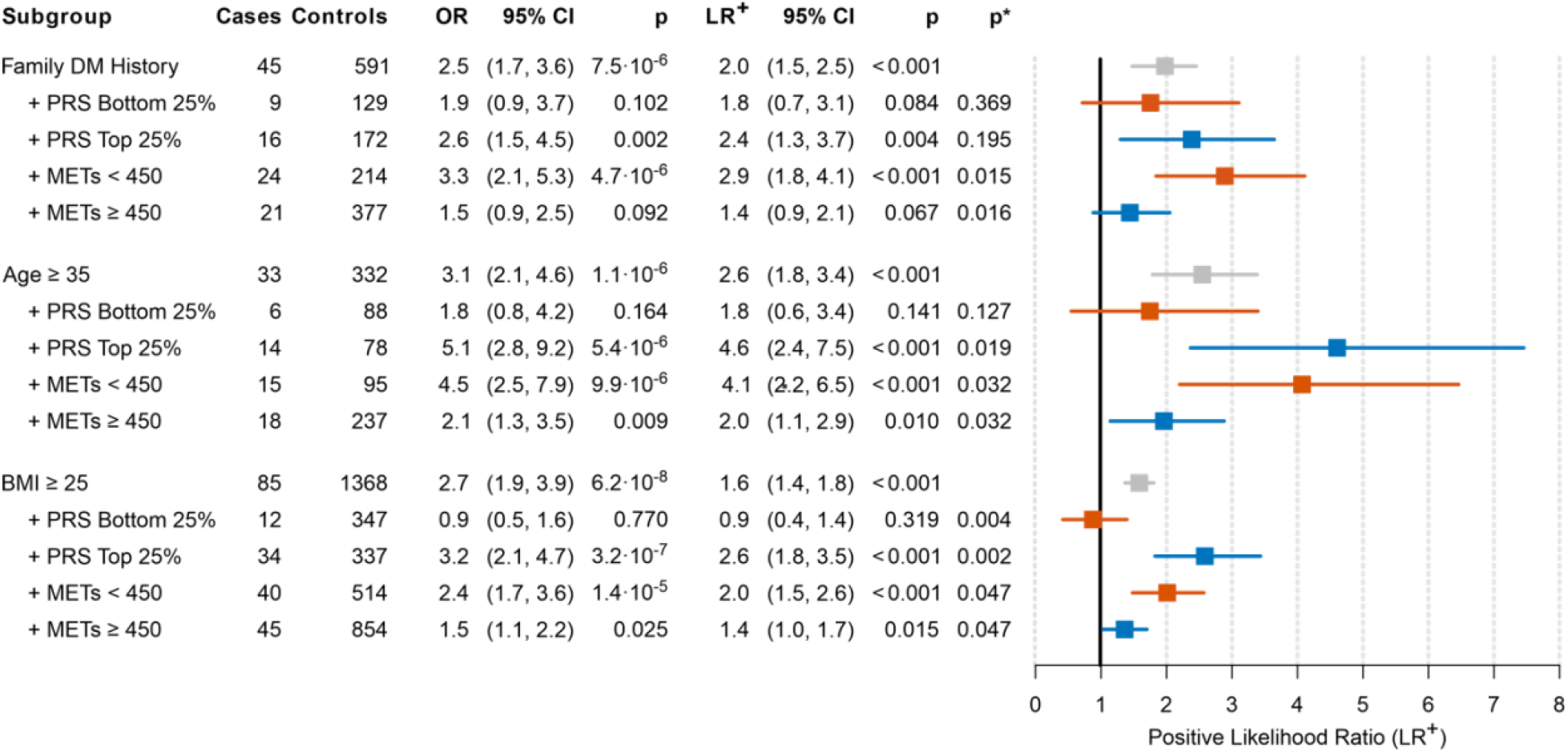
Influence of PRS and METs on GDM risk in the context of key clinical covariates (Family DM History, age, and BMI). The cases and controls list the number of participants in a subgroup on the left. The OR and LR^+^ values reflect the risk of developing GDM among subgroup participants with the rest of the cohort used as the reference group for OR and the entire cohort for LR^+^. OR p-value (*p*) was calculated using Fisher’s exact test. LR^+^ p-value (*p*) is the bootstrapped p-value of the LR^+^, where the reference group is all participants. LR^+^ p-value against parent subgroup (*p*\*) is the bootstrapped p-value of the LR^+^, where the reference group is the parent subgroup only. For example, among participants with BMI ≥ 25 (LR^+^ = 1.6), those with PRS in the bottom 25% have a significantly reduced GDM risk (LR^+^ = 0.9), with a p-value of 0.004. Furthermore, there is no statistical support that this subgroup (BMI ≥ 25 and PRS in the bottom 25%) differs from the entire cohort (p-value of 0.319).

In participants with age ≥ 35, we found that both high PRS (OR = 5.1) and low METs (OR = 4.5) were associated with increased odds of GDM diagnosis compared to the rest of the cohort. When the analysis was restricted to the parent group of participants with age ≥ 35 (LR^+^ = 2.6), we found that the subgroup with high PRS (LR^+^ = 4.6; *p*\* = 0.019) as well as the subgroup with low METs (LR^+^ = 4.1; *p*\* = 0.032) had further increased odds of GDM diagnosis. As with family history, the odds of GDM diagnosis were reduced for the participants with low PRS (LR^+^ = 1.8; *p*\* = 0.127) and high METs (LR^+^ = 2.0; *p*\* = 0.032).

In participants with BMI ≥ 25, we similarly found that both high PRS (OR = 3.2) and low METs (OR = 2.4) reflect increased odds for GDM compared to their respective remainders of the cohort. Among the participants with BMI ≥ 25 (LR^+^ = 1.6), those with high PRS (LR^+^ = 2.6; *p*\* = 0.002) and those with low METs (LR^+^ = 2.0; *p*\* = 0.047) were both found to have increased odds of GDM diagnosis. The subgroups of participants with low PRS (LR^+^ = 0.9; *p*\* = 0.004) and those with high METs (LR^+^ = 1.4; *p*\* = 0.047) exhibited reduced odds of GDM diagnosis. Interestingly, the participants with BMI ≥ 25 who had low PRS scores, had baseline-level odds of GDM diagnosis (Figure 3).

Next, we examined the interaction between PRS and METs related to the odds of developing GDM (Figure 4). Participants with high PRS and low METs show significantly higher odds of GDM (OR = 3.5) in comparison to the remaining participants. Even compared to the participants with high PRS (LR^+^ = 1.7) or participants with low METs (LR^+^ = 1.4), the participants who had both high PRS and low METs had considerably increased odds of GDM diagnosis (LR^+^ = 2.9, *p*\*<0.001). In contrast, participants with high PRS and high METs exhibit significantly lower LR^+^ compared to the parent subgroup of participants with high PRS (LR^+^ = 1.1, *p*\*<0.001) and the odds of GDM diagnosis similar to the baseline (*p* = 0.292). Finally, participants with low PRS and high METs had a significantly reduced risk of GDM diagnosis (OR = 0.5) compared to the participants outside of this group.

**Figure 4.**
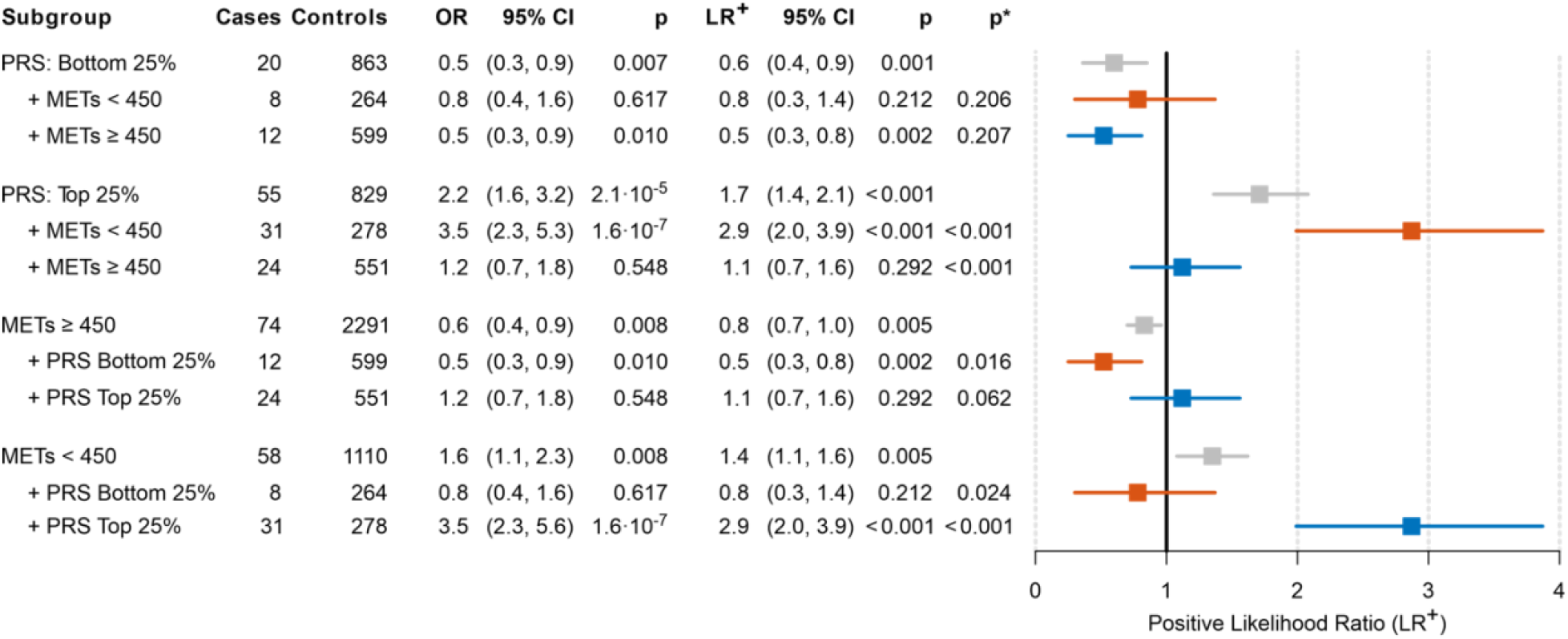
Cooperative effects of PRS and METs on GDM risk. The cases and controls list the number of participants in a subgroup on the left. The OR and LR^+^ values reflect the risk of developing GDM among subgroup participants with the rest of the cohort used as the reference group for OR and the entire cohort for LR^+^. LR^+^ p-value (*p*) is the bootstrapped p-value of the LR^+^, where the reference group is all participants. OR p-value (*p*) was calculated using Fisher’s exact test. LR^+^ p-value against parent subgroup (*p*\*) is the bootstrapped p-value of the LR^+^, where the reference group is the parent subgroup only. For example, among participants with METs<450 (LR^+^ = 1.4), those with PRS in the top 25% have a significantly increased GDM risk (LR^+^ = 2.9), with a p-value below 0.001. Furthermore, this subgroup (METs<450 and PRS in the top 25%) also has significantly higher risk from the entire cohort (p-value<0.001).

A formal test for the interaction between PRS and METs was carried out using the logit model as described in Methods. We estimate that *a* = −3.5, *b* = 0.42, *c* = 0.12, and *d* = 0.82, with the significance level for the interaction parameter *d* estimated as *p* = 0.028. Using an ordinary least-squares model, instead of the logit model, gave an even stronger support for the interaction between PRS and METs (*p*<0.001). We therefore conclude that our data provides support for the nonadditive relationship between the two covariates.

Finally, we evaluated the ability of machine learning models to predict GDM on the full cohort (baseline AUC = 0.710±0.042) using data from the first study visit only. The inclusion of PRS as a feature yields modest improvement in predictive performance compared to the baseline GDM prediction model (baseline + PRS AUC = 0.734±0.041). The addition of METs led to a similarly modest increase in performance (baseline + METs AUC = 0.708±0.046; baseline + PRS + METs AUC = 0.728±0.045).

## Discussion

We evaluated the influence of two critical factors, genetic predisposition and physical activity, on risk of GDM among self-reported White nulliparas with inferred European ancestry. We found that increased physical activity was associated with decreased risk of GDM, and this reduction in risk was particularly significant in individuals who were genetically predisposed to diabetes through PRS or family history. These results suggest a nonadditive relationship between genetic predisposition and physical activity. Physically active participants were overall at lower risk for GDM compared to other participants, regardless of PRS status. Similarly, participants with low PRS are at lower risk for GDM compared to other participants, regardless of activity level. These insights are consistent with existing evidence on the importance of lifestyle factors in risk of GDM^27^ and provides support for exercise interventions to improve pregnancy outcomes.^28^

Compared to previous studies on GDM risk, this work focuses exclusively on nulliparas. While previous work has found that GDM and other covariates from prior pregnancy are highly predictive for GDM risk, because all participants were nulliparous, this information was not available for our cohort. Further, the prediction of GDM risk in first pregnancies has remained an ongoing challenge despite the broader availability of informative factors such as BMI, age, and family history. Previously, Kawai et al.^14^ assessed the utility of PRS generated from loci associated with T2DM in GDM prediction and found a modest increase in predictive performance. Lamri et al.^29^ reported similar conclusions in a South Asian birth cohort, highlighting the potential for increased prediction accuracy when both PRS and other known GDM risk factors are considered. In the present study, we characterized the excess risk for GDM that may be attributable to risk factors including physical activity and PRS, both individually and jointly.

While both high PRS and low physical activity have been shown to independently increase the risk for GDM, the relationship between these two covariates is particularly valuable in clinical settings. Participants with high PRS and moderate-to-high activity levels in early pregnancy (METs≥450) exhibit similar GDM risk compared to the general population. Increased physical activity recommendations for genetically predisposed patients may serve to ameliorate some of the excess GDM risk. Further, these findings suggest the utility of PRS to stratify high-risk patients, which can be subject to more targeted interventions to mitigate modifiable lifestyle risk factors at the appropriate stages of pregnancy.

A strength of the work is the identification of the relationship between risk factors exhibited by patients that are most at-risk for GDM. While the implementation of PRS in clinical practice needs expansion to more diverse populations and careful evaluation prospectively in intervention trials, these findings suggest that targeted interventions could reduce adverse perinatal outcomes among more vulnerable patients. Furthermore, if patients with higher genetic risk constitute more clinically severe cases, then this could allow for improved understanding of the pathophysiological pathways involved and better elucidate the mechanisms of risk in GDM.

## Data Availability

All data produced in the present study will be made available online after peer review, and are available upon reasonable request to the authors.

## Limitations

First, the application of PRS with T2DM markers was used for GDM. Despite the reported similarities in genetic background between conditions, this may reduce the strength of our findings. The application of this PRS may suffer due to significant variability in effect size for loci between the two conditions, as well as differences in the socioeconomic and genetic background of these patients in comparison to the GWAS cohort used to derive the PRS.

Second, the PRS was calculated based upon GWAS that comprised European participants. Inclusion of SNVs from multi-ethnic GWAS, or GWAS from other population groups would demonstrate the effectiveness and generalizability of these metrics across other ethnic groups.

Third, the baseline features include quantitative and self-reported measurements that may have introduced uncertainty. Participants were selected for inclusion based upon computed race/ethnicity. However, replication of these methods with either self-reported race or inferred ethnicity returns conclusions that are largely similar (Supplementary Figures S2-S5).

Finally, as many phenotypes have shown that relative risk varies with age, differences in age may account for differences in PRS performance when applied to cohorts of different demographic makeup.^30^ For diseases where genetic relative risk decreases over age, genetic risk factors have stronger explanatory power among younger populations, compared to older ones.

## Conclusions

The risk of GDM diagnosis increases significantly for individuals with high PRS as well as those with low level of physical activity. Physical activity in early pregnancy is associated with reduced risk of GDM and reversal of the excess risk in individuals with strong genetic predisposition to GDM. Collectively, the findings of this work highlight the potential for targeted interventions that increase physical activity to mitigate GDM risk among nulliparous individuals who are at high risk through genetic predisposition, age, and family history of diabetes.

## Article Information

### Author Contributions

Drs. Haas and Radivojac had full access to all of the data in the study and take responsibility for the integrity of the data and the accuracy of the data analysis. Dr. Kymberleigh A. Pagel, Hoyin Chu, and Rashika Ramola contributed equally to this work as first authors.

Study concept and design: Radivojac, Haas, all authors

Acquisition of data: Guerrero, Ramola, Chu

Analysis and interpretation of data: Chu, Ramola, Pagel, Guerrero

Drafting of the manuscript: Pagel, Ramola, Chu

Critical revision of the manuscript for important intellectual content: Chung, Guerrero, Haas, Hahn, Natarajan, Parry, Radivojac, Reddy, Steller, Silver, Wapner, Yee

Statistical analysis: Chu, Ramola, Guerrero

Obtained funding: Haas, Hahn, Natarajan, Radivojac

Study supervision: Haas, Natarajan, Radivojac

## Conflict of Interest Disclosures

None reported.

## Funding/Support

Precision Health Initiative of Indiana University, National Institutes of Health award R01HD101246 to DMH, SN and PR. Cooperative agreement funding from the National Heart, Lung, and Blood Institute and the Eunice Kennedy Shriver National Institute of Child Health and Human Development: grant U10-HL119991 to RTI International; grant U10-HL119989 to Case Western Reserve University; grant U10-HL120034 to Columbia University; grant U10-HL119990 to Indiana University; grant U10-HL120006 to the University of Pittsburgh; grant U10-HL119992 to Northwestern University; grant U10-HL120019 to the University of California, Irvine; grant U10-HL119993 to University of Pennsylvania; and grant U10-HL120018 to the University of Utah. National Center for Research Resources and the National Center for Advancing Translational Sciences, National Institutes of Health to Clinical and Translational Science Institutes at Indiana University (grant UL1TR001108) and University of California, Irvine (grant UL1TR000153).

### Role of the Sponsors

The funders had no role in the design and conduct of the study; collection, management, analysis, and interpretation of the data; preparation, review, or approval of the manuscript; and decision to submit the manuscript for publication.

## Notes

### Competing Interest Statement

The authors have declared no competing interest.

### Author Declarations

All activities were covered by IRB at Indiana University. nuMoM2b was approved at IU as protocol Study number 1008-08 on 9/28/2010.

