## Supplement for "The influence of genetic predisposition and physical activity on risk of Gestational Diabetes Mellitus in the nuMoM2b cohort"

### Supplementary Materials

This document gives supplementary information for the manuscript titled "The influence of genetic predisposition and physical activity on risk of Gestational Diabetes Mellitus in the nuMoM2b cohort" by Pagel, Chu, Ramola, *et al*., 2022.

#### Selection of METs thresholds

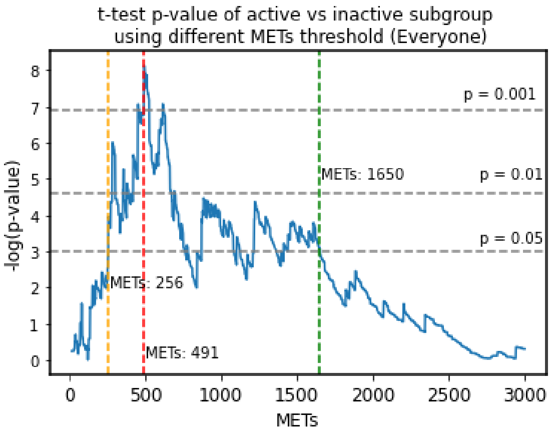


**Supplementary Figure S1.** The p-value of GDM incidence for the METs-based subgroups. The cohort was split based on the METs threshold shown on the x-axis and the two-sample t-test p-value, shown on the y-axis, was calculated using the binary vectors of the incidence of GDM in each group (1 = cases, 0 = controls). The yellow (METs = 256) and green (METs = 1650) dashed lines show the lowest and the largest METs value with a p-value below 0.05. The red dashed line shows the METs value (METs = 491) with the strongest separation between the two groups, based on the p-value.

#### Inferred European Participants

| Subgroup | Cases | Controls | OR | 95% CI | P | $LR^+$ | 95% CI | P | P* |
|---|---|---|---|---|---|---|---|---|---|
| Family DM History | 47 | 631 | 2.3 | (1.6, 3.3) | $1.91 \cdot 10^{-5}$ | 1.9 | (1.4, 2.3) | <0.001 | |
| +PRS Bottom 25% | 10 | 138 | 1.9 | (1.0, 3.7) | 0.075 | 1.8 | (0.8, 3.1) | 0.083 | 0.420 |
| +PRS Top 25% | 16 | 182 | 2.4 | (1.4, 4.1) | 0.004 | 2.2 | (1.2, 3.3) | 0.008 | 0.223 |
| +METs<450 | 25 | 229 | 3.1 | (2.0, 4.9) | $7.66 \cdot 10^{-6}$ | 2.7 | (1.8, 3.8) | <0.001 | 0.015 |
| +METs≥450 | 22 | 402 | 1.4 | (0.9, 2.3) | 0.138 | 1.4 | (0.9, 1.9) | 0.087 | 0.015 |
| Age>35 | 37 | 346 | 3.3 | (2.2, 4.8) | $4.51 \cdot 10^{-8}$ | 2.7 | (2.0, 3.5) | <0.001 | |
| +PRS Bottom 25% | 7 | 92 | 2.0 | (0.9, 4.3) | 0.103 | 1.9 | (0.6, 3.5) | 0.118 | 0.103 |
| +PRS Top 25% | 15 | 83 | 5.0 | (2.8, 8.8) | $3.20 \cdot 10^{-6}$ | 4.5 | (2.5, 7.2) | <0.001 | 0.025 |
| +METs<450 | 17 | 101 | 4.7 | (2.7, 8.0) | $1.62 \cdot 10^{-6}$ | 4.2 | (2.4, 6.6) | <0.001 | 0.025 |
| +METs≥450 | 20 | 245 | 2.2 | (1.4, 3.6) | 0.003 | 2.0 | (1.3, 3.0) | 0.003 | 0.025 |
| BMI>25 | 93 | 1455 | 2.7 | (1.9, 3.9) | $1.01 \cdot 10^{-8}$ | 1.6 | (1.4, 1.8) | <0.001 | |
| +PRS Bottom 25% | 15 | 370 | 1.0 | (0.6, 1.8) | 0.889 | 1.0 | (0.6, 1.5) | <0.001 | 0.009 |
| +PRS Top 25% | 37 | 353 | 3.2 | (2.2, 4.7) | $7.34 \cdot 10^{-8}$ | 2.6 | (1.9, 3.5) | <0.001 | 0.002 |
| +METs<450 | 42 | 552 | 2.3 | (1.6, 3.3) | $3.41 \cdot 10^{-5}$ | 1.9 | (1.4, 2.4) | <0.001 | 0.079 |
| +METs≥450 | 51 | 903 | 1.6 | (1.2, 2.3) | 0.006 | 1.4 | (1.1, 1.8) | 0.003 | 0.078 |

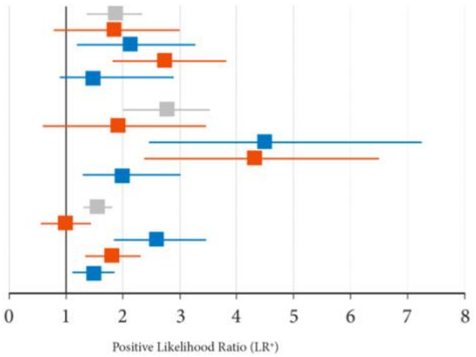


**Supplementary Figure S2.** Influence of PRS and METs on the GDM risk in the context of key clinical covariates (Family DM History, age, and BMI). The cases and controls list the number of participants in a subgroup on the left. The OR and $LR^+$ values reflect the risk of developing GDM among subgroup participants with the rest of the cohort used as the reference group for OR and the entire cohort for $LR^+$. OR p-value (*P*) was determined using Fisher's exact test. $LR^+$ p-value (*P*) is the bootstrapped p-value of the $LR^+$, where the reference group is all participants. $LR^+$ p-value against parent subgroup (*P**) is the bootstrapped p-value of the $LR^+$, where the reference group is the parent subgroup only.

| Subgroup | Cases | Controls | OR | 95% CI | P | $LR^+$ | 95% CI | P | P* |
|---|---|---|---|---|---|---|---|---|---|
| PRS Bottom 25% | 23 | 910 | 0.6 | (0.4, 0.9) | 0.010 | 0.6 | (0.4, 0.9) | 0.002 | |
| +METs<450 | 9 | 287 | 0.8 | (0.4, 1.5) | 0.634 | 0.8 | (0.3, 1.3) | 0.186 | 0.216 |
| +METs≥450 | 14 | 623 | 0.5 | (0.3, 0.9) | 0.017 | 0.6 | (0.3, 0.8) | 0.002 | 0.217 |
| PRS Top 25% | 60 | 874 | 2.2 | (1.6, 3.2) | $7.02 \cdot 10^{-6}$ | 1.7 | (1.4, 2.1) | <0.001 | |
| +METs<450 | 33 | 297 | 3.3 | (2.2, 5.0) | $1.29 \cdot 10^{-7}$ | 2.8 | (2.0, 3.8) | <0.001 | <0.001 |
| +METs≥450 | 27 | 577 | 1.2 | (0.8, 1.9) | 0.356 | 1.2 | (0.8, 1.6) | 0.189 | <0.001 |
| METs≥450 | 82 | 2405 | 0.7 | (0.5, 0.9) | 0.018 | 0.9 | (0.7, 1.0) | 0.016 | 0.016 |
| +PRS Bottom 25% | 14 | 623 | 0.5 | (0.3, 0.9) | 0.017 | 0.6 | (0.3, 0.8) | 0.002 | 0.016 |
| +PRS Top 25% | 27 | 577 | 1.2 | (0.8, 1.9) | 0.356 | 1.2 | (0.8, 1.6) | 0.189 | 0.051 |
| METs<450 | 61 | 1185 | 1.5 | (1.1, 2.1) | 0.018 | 1.3 | (1.0, 1.5) | 0.016 | 0.016 |
| +PRS Bottom 25% | 9 | 287 | 0.8 | (0.4, 1.5) | 0.634 | 0.8 | (0.3, 1.3) | 0.186 | 0.019 |
| +PRS Top 25% | 33 | 297 | 3.3 | (2.2, 5.0) | $1.29 \cdot 10^{-7}$ | 2.8 | (2.0, 3.8) | <0.001 | <0.001 |

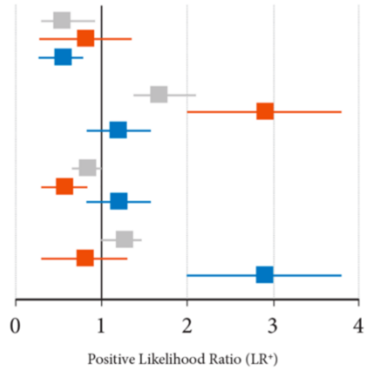


**Supplementary Figure S3.** Cooperative effects of PRS and METs on GDM risk. The cases and controls list the number of participants in a subgroup on the left. The OR and $LR^+$ values reflect the risk of developing GDM among subgroup participants with the rest of the cohort used as the reference group for OR and the entire cohort for $LR^+$. OR p-value (*P*) was determined using Fisher's exact test. $LR^+$ p-value (*P*) is the bootstrapped p-value of the $LR^+$, where the reference group is all participants. $LR^+$ p-value against parent subgroup (*P**) is the bootstrapped p-value of the $LR^+$, where the reference group is the parent subgroup only.

#### Self-Reported White Participants

| Subgroup | Cases | Controls | OR | 95% CI | P | $LR^+$ | 95% CI | P | P* |
|---|---|---|---|---|---|---|---|---|---|
| Family DM History | 47 | 663 | 2.4 | (1.7, 3.4) | $9.19 \cdot 10^{-6}$ | 1.9 | (1.5, 2.4) | <0.001 | |
| +PRS Bottom 25% | 9 | 145 | 1.7 | (0.9, 3.4) | 0.122 | 1.7 | (0.7, 2.9) | 0.097 | 0.319 |
| +PRS Top 25% | 17 | 195 | 2.5 | (1.5, 4.3) | 0.002 | 2.3 | (1.3, 3.5) | 0.028 | 0.175 |
| +METs<450 | 24 | 235 | 3.1 | (2.0, 4.9) | $1.09 \cdot 10^{-5}$ | 2.7 | (1.8, 3.9) | 0.001 | 0.018 |
| +METs≥450 | 23 | 428 | 1.5 | (1.0, 2.4) | 0.078 | 1.4 | (0.9, 2.0) | 0.054 | 0.018 |
| Age>35 | 34 | 376 | 2.9 | (1.9, 4.3) | $1.61 \cdot 10^{-6}$ | 2.4 | (1.7, 3.2) | <0.001 | |
| +PRS Bottom 25% | 6 | 99 | 1.7 | (0.7, 3.8) | 0.276 | 1.6 | (0.5, 3.2) | 0.170 | 0.119 |
| +PRS Top 25% | 14 | 86 | 4.8 | (2.6, 8.6) | $9.47 \cdot 10^{-6}$ | 4.4 | (2.3, 7.1) | <0.001 | 0.017 |
| +METs<450 | 15 | 104 | 4.2 | (2.4, 7.5) | $1.66 \cdot 10^{-5}$ | 3.9 | (2.1, 6.2) | <0.001 | 0.033 |
| +METs≥450 | 19 | 272 | 2.0 | (1.2, 3.3) | 0.008 | 1.9 | (1.1, 2.8) | 0.012 | 0.033 |
| BMI>25 | 91 | 1497 | 2.8 | (2.0, 4.0) | $3.58 \cdot 10^{-9}$ | 1.6 | (1.4, 1.8) | <0.001 | |
| +PRS Bottom 25% | 13 | 377 | 0.9 | (0.5, 1.6) | 0.886 | 0.9 | (0.5, 1.5) | 0.379 | 0.004 |
| +PRS Top 25% | 36 | 371 | 3.2 | (2.1, 4.7) | $1.27 \cdot 10^{-7}$ | 2.6 | (1.9, 3.4) | <0.001 | 0.002 |
| +METs<450 | 42 | 551 | 2.5 | (1.7, 3.6) | $5.70 \cdot 10^{-6}$ | 2.0 | (1.5, 2.6) | <0.001 | 0.043 |
| +METs≥450 | 49 | 946 | 1.6 | (1.1, 2.3) | 0.010 | 1.4 | (1.1, 1.7) | 0.007 | 0.043 |

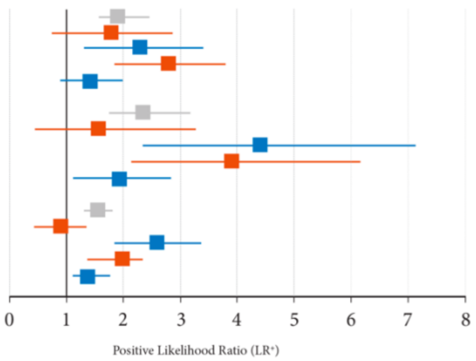


**Supplementary Figure S4.** Influence of PRS and METs on the GDM risk in the context of key clinical covariates (Family DM History, age, and BMI). The cases and controls list the number of participants in a subgroup on the left. The OR and $LR^+$ values reflect the risk of developing GDM among subgroup participants with the rest of the cohort used as the reference group for OR and the entire cohort for $LR^+$. OR p-value (*P*) was determined using Fisher's exact test. $LR^+$ p-value (*P*) is the bootstrapped p-value of the $LR^+$, where the reference group is all participants. $LR^+$ p-value against parent subgroup (*P**) is the bootstrapped p-value of the $LR^+$, where the reference group is the parent subgroup only.

| Subgroup | Cases | Controls | OR | 95% CI | P | $LR^+$ | 95% CI | P | P* |
|---|---|---|---|---|---|---|---|---|---|
| PRS Bottom 25% | 21 | 948 | 0.5 | (0.3, 0.8) | 0.005 | 0.6 | (0.4, 0.8) | 0.001 | |
| +METs<450 | 9 | 288 | 0.8 | (0.4, 1.6) | 0.744 | 0.8 | (0.4, 1.5) | 0.286 | 0.129 |
| +METs≥450 | 12 | 660 | 0.4 | (0.2, 0.8) | 0.004 | 0.5 | (0.2, 0.8) | <0.001 | 0.129 |
| PRS Top 25% | 57 | 913 | 2.2 | (1.5, 3.0) | $2.39 \cdot 10^{-5}$ | 1.7 | (1.3, 2.0) | <0.001 | |
| +METs<450 | 32 | 302 | 3.4 | (2.3, 5.1) | $1.20 \cdot 10^{-7}$ | 2.8 | (2.0, 3.8) | <0.001 | <0.001 |
| +METs≥450 | 25 | 611 | 1.1 | (0.7, 1.7) | 0.640 | 1.1 | (0.7, 1.5) | 0.313 | <0.001 |
| METs≥450 | 79 | 2542 | 0.6 | (0.4, 0.9) | 0.007 | 0.8 | (0.7, 1.0) | 0.005 | |
| +PRS Bottom 25% | 12 | 660 | 0.4 | (0.2, 0.8) | 0.004 | 0.5 | (0.2, 0.8) | <0.001 | 0.007 |
| +PRS Top 25% | 25 | 611 | 1.1 | (0.7, 1.7) | 0.640 | 1.1 | (0.7, 1.5) | 0.313 | 0.073 |
| METs<450 | 60 | 1195 | 1.6 | (1.1, 2.3) | 0.007 | 1.3 | (1.1, 1.6) | 0.005 | |
| +PRS Bottom 25% | 9 | 288 | 0.8 | (0.4, 1.6) | 0.744 | 0.8 | (0.4, 1.5) | 0.281 | 0.039 |
| +PRS Top 25% | 32 | 302 | 3.4 | (2.3, 5.1) | $1.20 \cdot 10^{-7}$ | 2.8 | (2.0, 3.8) | <0.001 | <0.001 |

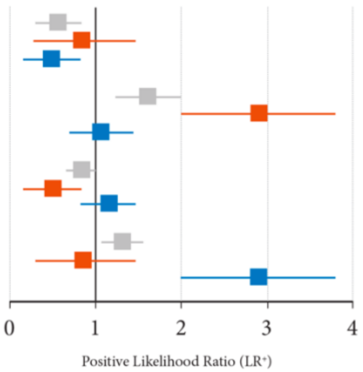


**Supplementary Figure S5.** Cooperative effects of PRS and METs on GDM risk. The cases and controls list the number of participants in a subgroup on the left. The OR and $LR^+$ values reflect the risk of developing GDM among subgroup participants with the rest of the cohort used as the reference group for OR and the entire cohort for $LR^+$. OR p-value (*P*) was determined using Fisher's exact test. $LR^+$ p-value (*P*) is the bootstrapped p-value of the $LR^+$, where the reference group is all participants. $LR^+$ p-value against parent subgroup (*P**) is the bootstrapped p-value of the $LR^+$, where the reference group is the parent subgroup only.
